# Public Acceptability of Self-Payment for Community Pharmacists’ Services: A National Online Survey

**DOI:** 10.64898/2026.11.15.26368139

**Authors:** Kimberly A. Pesaturo, John-Henry Pezzuto, Melissa Mattison, John M. Pezzuto

**Author notes:** To whom correspondence should be addressed: Kimberly A. Pesaturo, College of Pharmacy and Health Sciences, Western New England University, 1215 Wilbraham Road, Springfield, MA, USA, 01119. These authors contributed equally to this work (Kimberly A. Pesaturo, John-Henry Pezzuto).

## Abstract

**Background:** The traditional business model for sustaining United States (U.S.) community pharmacies has relied on income generated by product sales. A fee-for-service model could help alleviate insufficient profit by monetizing conventionally free pharmacy services if patients are willing to pay out-of-pocket.

**Objectives:** This study evaluated public acceptability of out-of-pocket payment for community pharmacist-provided services, variation in payment acceptability across services, and assessed whether payment acceptability differed by respondent characteristics and trust constructs.

**Methods:** This study represents a cross-sectional online survey of U.S. adults conducted during spring 2026 using a non-probability sampling approach. Primary outcomes were binary stated payment-acceptability judgments for a variety of pharmacist and health services. The survey used a between-subjects randomized payment-framing design, where participants were assigned to either a stated-price condition or an any-payment condition. Secondary outcomes included trust designations and participant demographic characteristics.

**Results:** A total of 1069 respondents completed the survey. A $20 fee was viewed more favorably than a general “any amount of money” fee. In the $20 stated-price condition, agreement was highest for osteoporosis screening (75.8%) and travel-related vaccines (69.0%), and lowest for antibiotic use consultation (23.9%) and wound care (32.1%). Older respondents and Hispanic respondents were less likely to view payment as acceptable, whereas men and respondents with greater pharmacist trust were more likely to view payment as acceptable.

**Conclusion:** Although patients historically have received many community pharmacy services at no cost, this study demonstrates that the concept of self-payment ifor certain services is considered acceptable, especially for services not typically provided.

**Key points:**

- In a quota-matched survey of 1,069 U.S. adults, this study evaluated public acceptability of direct out-of-pocket payment for 27 community pharmacist-provided clinical services.
- Public acceptability of a $20 out-of-pocket payment varied substantially by service, ranging from 23.9% for antibiotic-use consultation to 75.8% for osteoporosis screening. Higher acceptability was observed for discrete or specialized services, including travel-related vaccines and pharmacogenomic result review.
- Payment acceptability was associated with respondent characteristics and trust in pharmacists. Older respondents and Hispanic respondents were less likely to view payment as acceptable, whereas men and respondents with greater pharmacist trust were more likely to view payment as acceptable.

## 1. Background

Around the turn of the 20^th^ century, the community pharmacist held a privileged position in United States (U.S.) society. Elixirs, tablets, creams, etc., were skillfully compounded and dispensed. The pharmacist played an important role in the healthcare system, being a trusted and readily accessible expert regarding various ailments, and in a prime position to provide professional advice and remedies. The profession reached an inflection point around 1920, dubbed the “Soda Fountain Era” by Urick and Meggs (1920-1949) [1]. The expectations for a practicing pharmacist during this period were elaborated by LaWall: The 1922 American Pharmaceutical Association (APhA) Code of Ethics, many of which remain valid today [2]. Noteworthy points include: “He [the pharmacist] should perform no act, nor should he be a party to any transaction which will bring discredit to his profession…; He should not allow his name to be used in connection to advertisements…; He should not undersell a fellow pharmacist for the sake of commercial advantage…; The Pharmacist should be willing to join in any constructive effort to promote the public welfare…”. However, regarding the relationship of the pharmacist to the physician, the 1922 Code written by LaWall was more stifling.^2^ Here, LaWall minimizes the pharmacists’ role in diagnostics and the provision of informed care.

A great deal has changed over the decades. However, remnants of the 1922 Code of Ethics [2] echoed during the recent opioid crisis, when some pharmacists, with limited access to patient medical records, filled opioid prescriptions written by DEA-registered physicians. Of course, had the code still applied, companies like CVS, Walgreens, and Walmart would not have been found liable and required to pay a major portion of opioid settlements valued in the billions of dollars [3]. This reinforces the current pharmacist role in safeguarding public health, inclusive of acting on prescriptions that ostensibly meet all legal requirements.

Herein lies an important segment of the problematic current day U.S. fiscal pharmacy crisis. Again, per LaWall: “The Pharmacist should be properly remunerated by the public for his knowledge and skill when used in its behalf in compounding prescriptions, and his fee for such professional work as well as the cost of the ingredients” [2]. It may be suggested, at least in large chain pharmacies, that the community pharmacist receives payment for knowledge and skill by the employer. But if the employer’s revenue is dependent on the product dispensed by the pharmacist, and that product is priced in a manner that is insufficient to cover costs, i.e., a negative profit margin, there is an incongruency that should be remedied.

Hope currently rests on passage and enactment of legislation such as the ECAPS Act (H.R. 3164; Equitable Community Access to Pharmacist Services Act) which would allow Medicare Part B reimbursement for specific pharmacist services [4]. Vaccinations, which are now commonplace at community pharmacies, yield around $40 per vaccine [5]. Presumably, other services of equal or even greater value for the public could be offered if adequate compensation were provided. In the meanwhile, however, responsibility for the well-being of patients rests on the shoulders of pharmacists, irrespective of payment.

Fiscal transformation, such as payment for services rendered through Medicare Part B, private insurance companies, or others, is necessary to enable many community pharmacists to practice and stay solvent. However, this solution remains in the political arena and is beyond the scope of our current proposition. We set out to explore services that can be provided by a community pharmacist that are encapsulated within the full scope of their license and are at times provided without a fee. We recognize that private citizens are accustomed to paying for goods and services they demand. But would the public be willing to pay for a beneficial service provided legally, proficiently, and conveniently at a local pharmacy by a community pharmacist?

Any such service would be subject to the concept of price elasticity. Therefore, we pondered services that might be considered somewhat analogous. For example, when obtaining a medical service, consumers are accustomed to being responsible for copayment. Similarly, acquiring a prescription drug normally requires an out-of-pocket expense, either in the form of a copayment or a balance due when using various pharmacy discount platforms. Accordingly, as we explored the concept of pay-for-service, we were mindful of limiting the fee levied by community pharmacists to remain within similar fiscal boundaries.

## 2. Objectives

This study evaluated public acceptability of direct out-of-pocket payment for community pharmacist-provided services in the United States. We examined variation in payment acceptability across services and assessed whether payment acceptability differed by respondent characteristics and trust in community pharmacists.

## 3. Methods

We conducted a preregistered cross-sectional online survey of U.S. adults during spring 2026. The primary outcomes were binary stated payment-acceptability judgments for a variety of pharmacist and health services. The survey used a between-subjects randomized payment-framing design, in which participants were assigned to either a stated-price condition or an any-payment condition. Secondary outcomes included trust designations regarding local community pharmacists and participant demographic characteristics. This study was approved by the Institutional Review Board of Western New England University and preregistered on AsPredicted (#283225). Replication materials for both studies are available in the Harvard Dataverse (https://doi.org/10.7910/DVN/ZFAVPQ).

### 3.1 Population

We recruited U.S. adults through Prolific, an internet-based participant recruitment platform, using a quota-matched nonprobability sampling approach [6]. The sample was quota matched to the U.S. adult population on gender, age, and race/ethnicity. The target sample size was selected to approximate U.S. population benchmarks while providing sufficient sample size to examine heterogeneity across demographic groups. Participants were eligible to participate if they were 18 years of age or older, proficient in English, and residing in the U.S.

### 3.2 Survey Instrument

The survey consisted of 27 binary payment-acceptability questions about clinical services that may be provided by a community pharmacist (Table 1). The choice of clinical services was guided by published literature describing common pharmacist-provided services and proposed charges for those services [7–10]. The survey was piloted internally by pharmacists, social scientists, and non-specialists to test question comprehension and completion time. Because many of these services may not have been directly relevant to all survey participants, we elicited judgments about whether payment would be acceptable for a *typical patient* paying out of pocket for each service, rather than participants’ own personal willingness to pay. To support informed responses, each question included a brief description of the clinical service being evaluated.

**Table 1.**
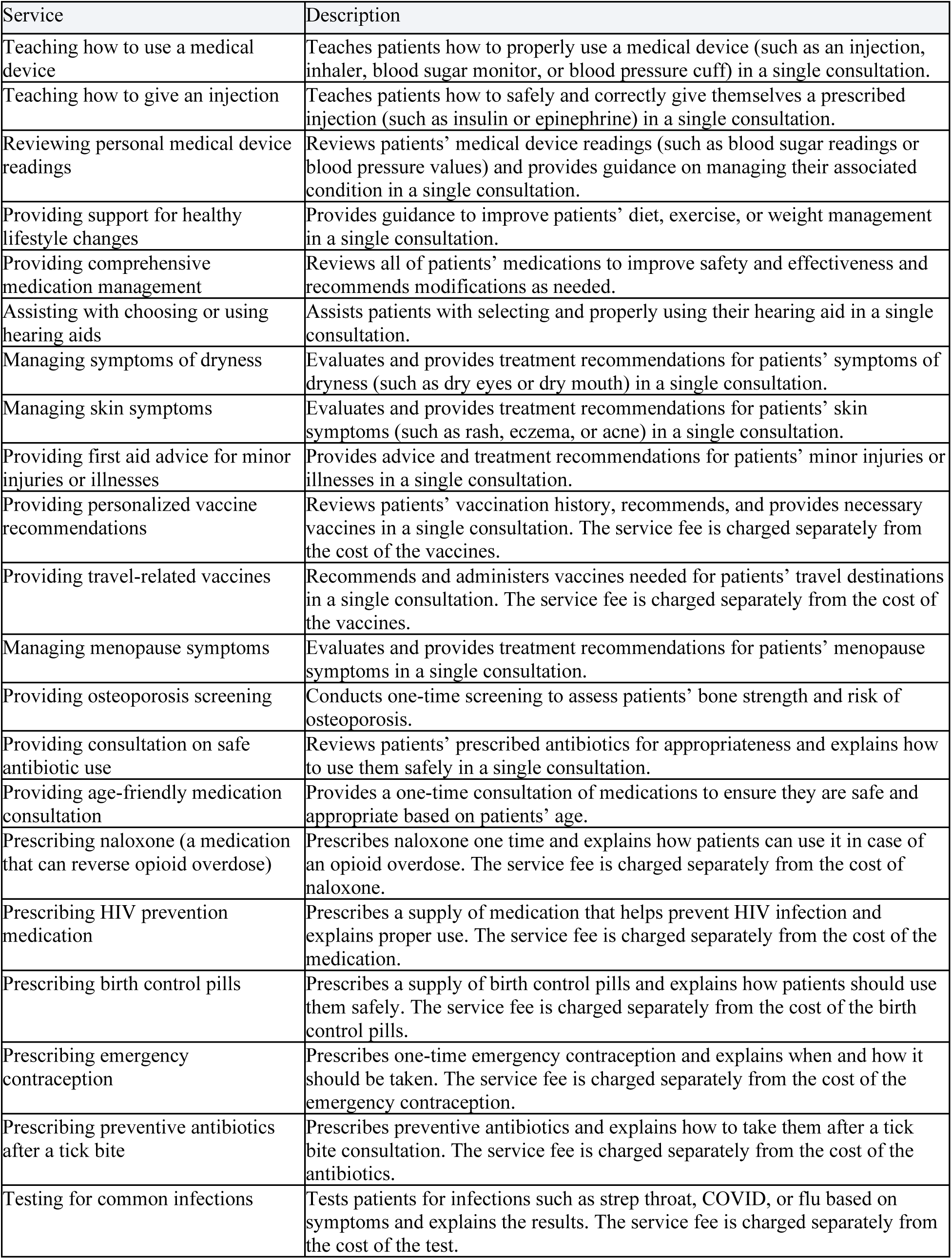
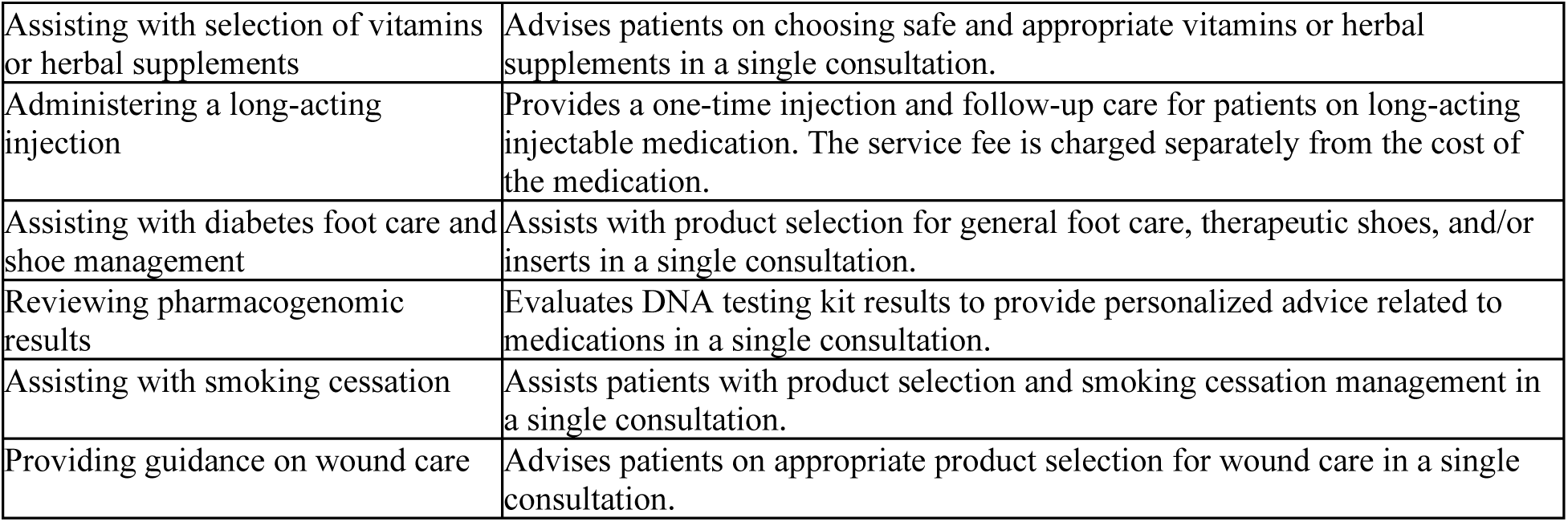
Survey services and descriptions.

Before beginning the payment-acceptability questions, participants were required to answer comprehension questions about the meaning of out-of-pocket payment and the fact that the questions referred to a typical patient rather than to themselves. Participants were not excluded for initially answering these comprehension questions incorrectly. Instead, participants were given as many attempts as needed to answer the questions correctly. This process accomplished the goal of assuring all participants began the survey with the same basic understanding of key determinates. Participants took an average of approximately 10 minutes to complete the survey (median: 8 minutes 45 seconds).

Payment framing was randomized at the participant level and remained fixed across all 27 service questions. In the $20 stated-price condition, participants were asked a yes/no question about whether it would be acceptable for a typical patient to pay $20 out of pocket for a given service. The $20 price point was chosen as a plausible lower-end out-of-pocket payment for pharmacist-provided services, recognizing that this amount is in the general range of an insurance co-payment that respondents may be accustomed to providing out-of-pocket. We applied the same price across all 27 services to facilitate comparisons of payment acceptability across services. We opted to use a binary choice elicitation to make the decisions simpler and easier to interpret for participants [11].

In the any-payment condition, participants were asked whether it would be acceptable for a typical patient to pay any amount of money for a given service. This condition was designed to assess the acceptability of out-of-pocket payment in principle, rather than whether payment was acceptable at a specific positive price point. Participants saw one clinical service question per page, and questions were presented in randomized order.

After completing the payment-acceptability questions, participants completed a follow-up survey. Participants first completed nine Likert-scale items assessing perceptions of trust in their local community pharmacist, with response options ranging from strongly disagree to strongly agree. These items were developed based on a systematic review that identified dimensions of trust in health professionals [12]. Participants then answered questions about personal demographics, geographic location, distance to the nearest pharmacy, timing of their last pharmacy visit, presence of chronic medical conditions, and self-designation as a healthcare professional. Please see Supplemental Figure 3.

### 3.3 Replication Study

We replicated our main study using a university student subject pool, where students participate for course credit. The replication study was not preregistered, and all students were eligible to participate. Students were recruited through the behavioral laboratory at the Rady School of Management at the University of California, San Diego. Of the 218 respondents, 143 (65.6%) identified as female and 75 (34.4%) as male. Race selections were Asian or Pacific Islander (141; 64.7%), Hispanic (44; 20.2%), white (43; 19.7%), other (10; 4.6%), Black (3; 1.4%), and Native American (1; 0.5%); respondents could select multiple options. Data collection ran in the lab for approximately one week from April 20 through April 24, 2026. All participants provided informed consent before participating.

Participants answered the same 27 payment-acceptability questions as in the main survey. They were randomly assigned at the respondent level to one of three payment-framing conditions: $20, or any amount of money, and a new $5 condition. We additionally included the $5 condition to address potential framing effects of quota-matched study, which we describe in greater detail in the results section. In total, there were 73 participants in the $20 condition, 73 in the $5 condition, and 72 in the any-amount condition. Unlike the main survey, we did not ask participants about their age, healthcare-professional status, marital status, education, household income, or the region where they currently lived in the follow-up questionnaire, because of the relatively homogenous sample population. Please see Supplemental Figure 4.

### 3.4 Statistical Analysis

Payment-acceptability responses were coded as 1 if participants indicated that payment would be acceptable and 0 otherwise. For each clinical service, we calculated the proportion of participants indicating that payment would be acceptable, separately by payment-framing condition. To create the pharmacist trust index, we averaged the nine trust items for each respondent after excluding items where the participant reported “no basis for observation.” Internal consistency was high in both samples (Cronbach’s α = .934 for Prolific and .936 for UCSD; complete-case n = 893 and 175, respectively). An exploratory factor analysis found that all nine items were strongly associated with a shared trust dimension in both samples (Prolific: .74–.91; UCSD: .74–.89), supporting their use in an overall trust score.

To examine heterogeneity in payment acceptability, we estimated linear probability models with payment acceptability as the dependent variable [13]. Models included participant characteristics, the pharmacist trust index, and payment-framing condition as predictors. Because each participant answered multiple service questions, standard errors were clustered at the participant level to account for within-participant correlation.

Statistical analyses were conducted in R version 4.5.3.

## 4. Results

A total of 1,069 respondents completed the Prolific survey and were included in the analysis. Demographics were balanced across experimental conditions. The sample included 532 female respondents, 518 male respondents, and 19 non-binary respondents. Respondents were distributed across age groups, including 14.6% ages 18-26, 14.4% ages 27-34, 25.3% ages 35-49, 35.8% ages 50-67, and 9.9% over age 67. Most respondents identified as Caucasian or white (63.0%), while 12.3% identified as African American or Black, 8.1% as Hispanic, 5.7% as Asian or Pacific Islander, 1.0% as Native American, 1.5% as other, and 8.3% as multiracial. A total of 84 respondents (7.9%) reported being healthcare professionals. Educational attainment was relatively high, with 81.8% of respondents reporting college or higher education. A full demographic profile is provided in Table 2.

**Table 2.**
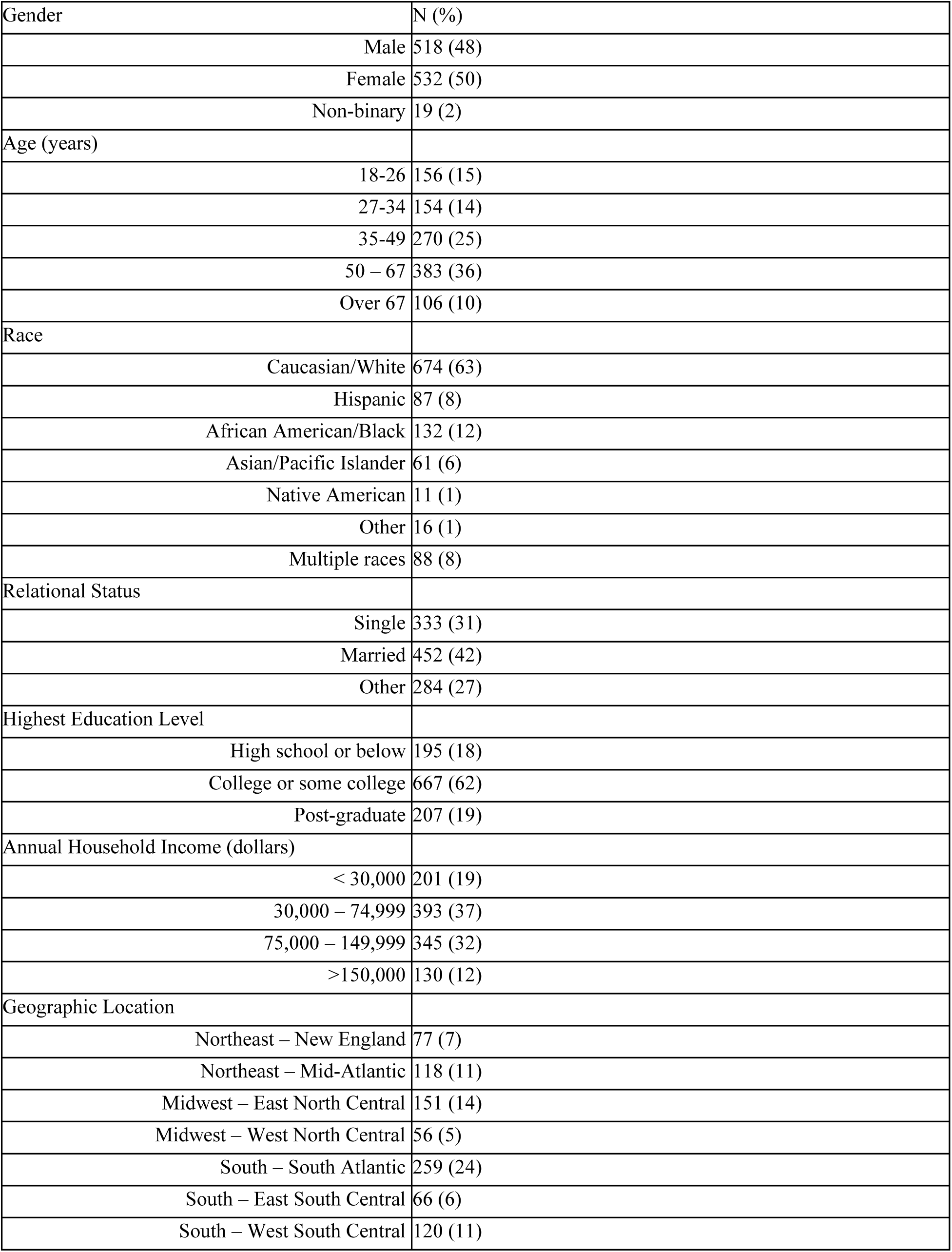
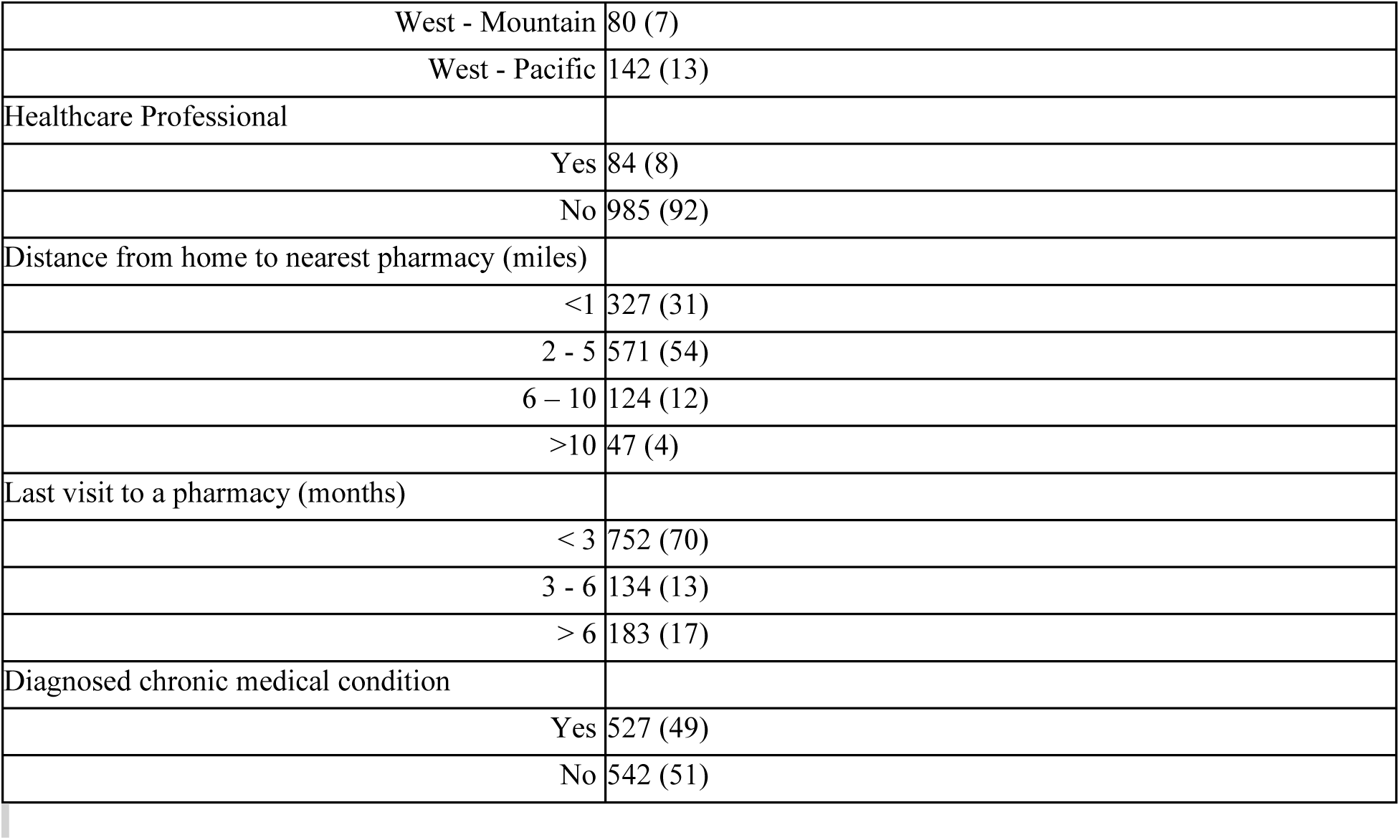
Demographics of survey respondents.

| Gender | N (%) |
|---|---|
| Male | 518 (48) |
| Female | 532 (50) |
| Non-binary | 19 (2) |
| Age (years) |  |
| 18-26 | 156 (15) |
| 27-34 | 154 (14) |
| 35-49 | 270 (25) |
| 50 – 67 | 383 (36) |
| Over 67 | 106 (10) |
| Race |  |
| Caucasian/White | 674 (63) |
| Hispanic | 87 (8) |
| African American/Black | 132 (12) |
| Asian/Pacific Islander | 61 (6) |
| Native American | 11 (1) |
| Other | 16 (1) |
| Multiple races | 88 (8) |
| Relational Status |  |
| Single | 333 (31) |
| Married | 452 (42) |
| Other | 284 (27) |
| Highest Education Level |  |
| High school or below | 195 (18) |
| College or some college | 667 (62) |
| Post-graduate | 207 (19) |
| Annual Household Income (dollars) |  |
| < 30,000 | 201 (19) |
| 30,000 – 74,999 | 393 (37) |
| 75,000 – 149,999 | 345 (32) |
| >150,000 | 130 (12) |
| Geographic Location |  |
| Northeast – New England | 77 (7) |
| Northeast – Mid-Atlantic | 118 (11) |
| Midwest – East North Central | 151 (14) |
| Midwest – West North Central | 56 (5) |
| South – South Atlantic | 259 (24) |
| South – East South Central | 66 (6) |
| South – West South Central | 120 (11) |
| West - Mountain | 80 (7) |
| West - Pacific | 142 (13) |
| Healthcare Professional |  |
| Yes | 84 (8) |
| No | 985 (92) |
| Distance from home to nearest pharmacy (miles) |  |
| <1 | 327 (31) |
| 2 - 5 | 571 (54) |
| 6 – 10 | 124 (12) |
| >10 | 47 (4) |
| Last visit to a pharmacy (months) |  |
| < 3 | 752 (70) |
| 3 - 6 | 134 (13) |
| > 6 | 183 (17) |
| Diagnosed chronic medical condition |  |
| Yes | 527 (49) |
| No | 542 (51) |

### 4.1 Public Acceptability of Paying Out-of-pocket

The main outcome was the proportion of respondents indicating that it would be acceptable for a typical patient to pay for each pharmacist-provided clinical service. Figure 1 summarizes these results. The left panel shows responses from participants randomly assigned to the $20 stated-price condition, and the right panel shows responses from participants randomly assigned to the any-payment condition. The main result from Figure 1 is that public acceptability of out-of-pocket payment varied substantially across pharmacist-provided services. In the $20 stated-price condition, agreement was highest for osteoporosis screening (75.8%), travel-related vaccines (69.0%), and reviewing pharmacogenomic results (68.8%). The services with the highest acceptability may be characterized as highly specialized services that require additional pharmacist expertise.

**Figure 1.**
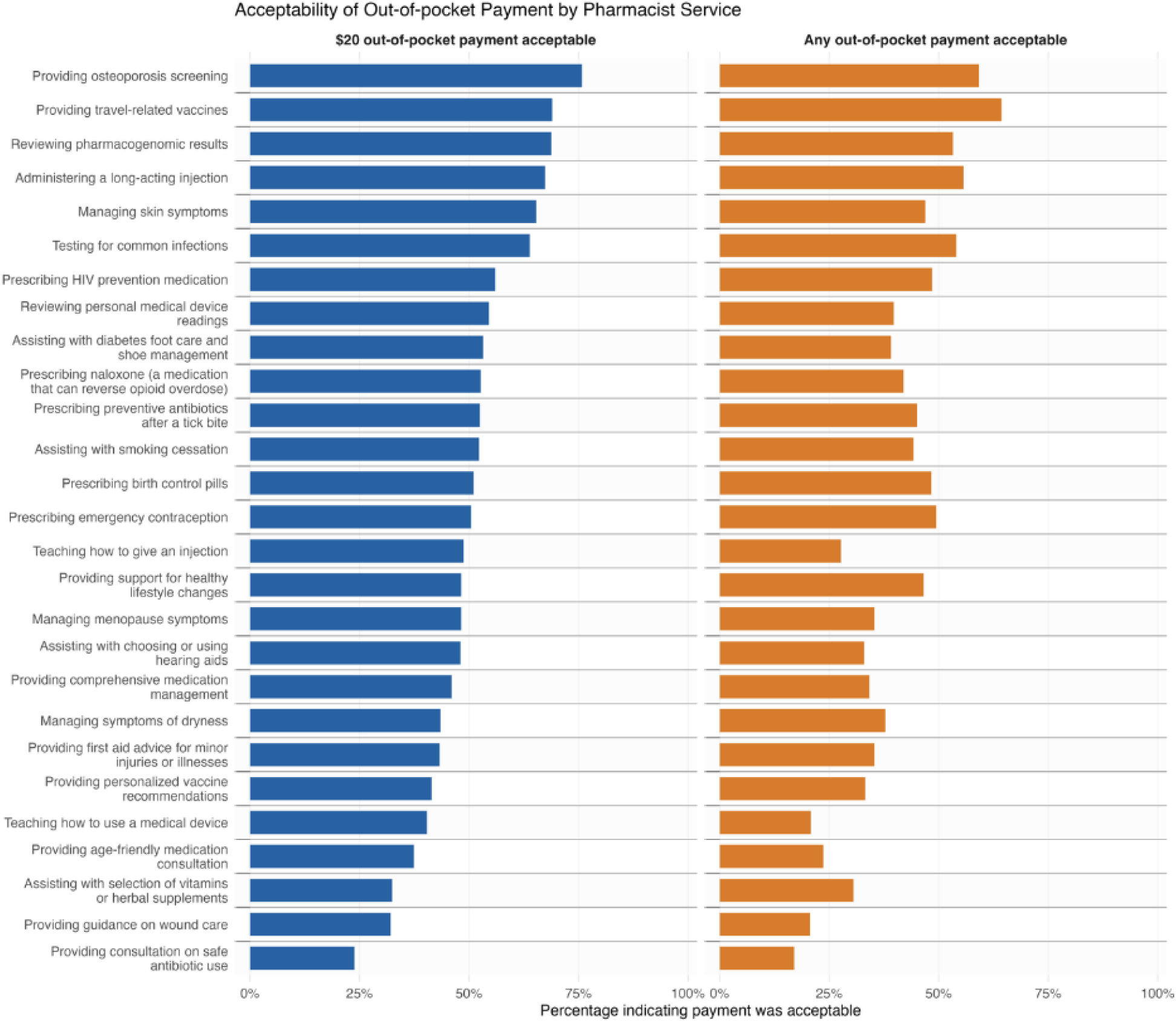
Acceptability of out-of-pocket payment by pharmacist service. The dependent variable is an indicator equal to 1 if the respondent agreed that the randomly assigned price was acceptable for the pharmacy service, and 0 otherwise. Respondents were randomly assigned to either the $20 condition or the anything-at-all condition. Bars show the mean of this indicator by service and condition. Services are sorted by the agreement rate in the $20 condition, from highest to lowest, and the same service order is used in both panels.

By contrast, agreement was lowest for services involving counseling, guidance, or support that respondents may view as already embedded in routine pharmacy practice or with information easily accessible online. Agreement was lowest for consultation on safe antibiotic use (23.9%), guidance on wound care (32.1%), and assistance with selection of vitamins or herbal supplements (32.5%). For these services, most respondents did not indicate that out-of-pocket payment would be acceptable.

Importantly, there was substantial heterogeneity in beliefs about payment acceptability even for the most highly endorsed services. In the $20 condition, approximately one-fourth of respondents did not view payment as acceptable for osteoporosis screening, and nearly one-third did not view payment as acceptable for travel-related vaccines or pharmacogenomic results review. This variation appears to reflect service-specific judgments rather than general opposition to out-of-pocket payment: 92.4% of respondents viewed payment as acceptable for some services but not others. Thus, while some pharmacist-provided services received meaningful public support, direct out-of-pocket payment did not receive broad consensus across services, and respondents differed in which services they viewed as appropriate candidates for direct payment. In the sections that follow, we examine which respondents are more likely to view out-of-pocket payment as acceptable and whether trust in pharmacists helps explain this variation.

### 4.2 Correlates of Public Acceptability

Understanding which respondents are more likely to view out-of-pocket payment as acceptable may help identify barriers to direct patient payment for pharmacist-provided services, as well as populations that may be more receptive to these services. Figure 2 examines whether payment acceptability varied systematically by respondents’ background characteristics and trust in community pharmacists. Points show estimated coefficients from linear probability models in which the dependent variable was whether the respondent indicated that payment would be acceptable for a given pharmacist-provided service. Models included service fixed effects and an indicator for payment-framing condition. All heterogeneity results are shown in Table 3. Rate of agreement to pay by service, condition, and gender are illustrated in Supplemental Figure 1.

**Figure 2.**
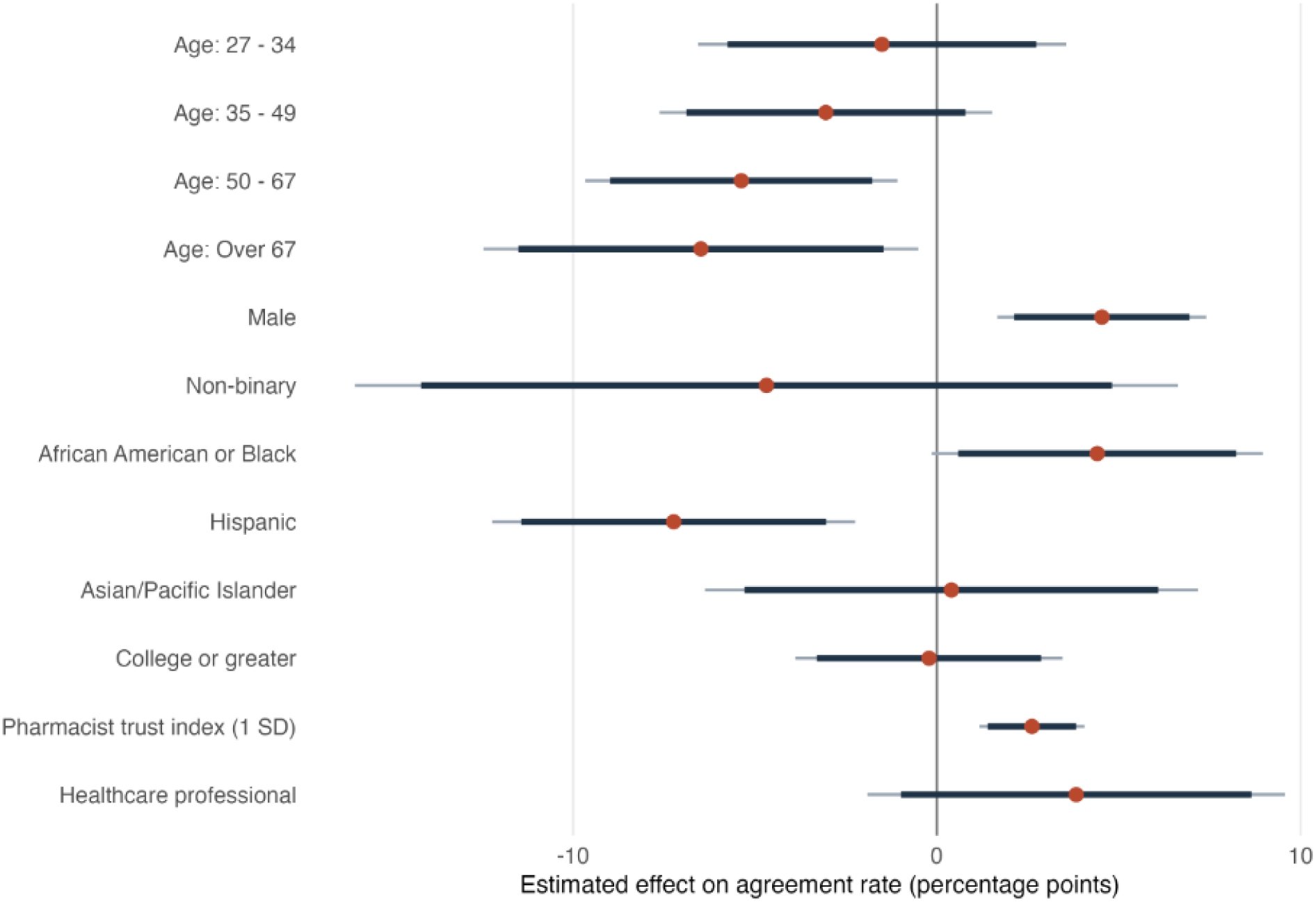
Correlates of payment acceptability. The outcome is an indicator for whether the respondent agreed to pay the posted price for a pharmacy service. Coefficients are from a linear probability model with service fixed effects and participant-clustered standard errors. The $20 condition is included as a control but not shown. Age coefficients are relative to respondents ages of 18-26. Male is relative to female respondents. Race/ethnicity indicators compare respondents who selected that category with white respondents. College or greater is relative to high school diploma or below. Healthcare professional is relative to not being a healthcare professional. The pharmacist trust index is the standardized average of the nine pharmacist-trust items. Points show coefficient estimates; bars show 90% and 95% confidence intervals.

**Table 3.** Heterogeneity results.

| **Characteristic** |  | **Estimate (pp)** | **SE (pp)** | **90% CI** | **95% CI** | ***p*-value** |
|---|---|---|---|---|---|---|
| Age: 27 - 34 |  | -1.5 | 2.6 | [-5.8, 2.7] | [-6.6, 3.6] | 0.55925 |
| Age: 35 - 49 |  | -3.1 | 2.3 | [-6.9, 0.8] | [-7.6, 1.5] | 0.19126 |
| Age: 50 - 67 |  | -5.4 | 2.2 | [-9.0, -1.8] | [-9.7, -1.1] | 0.01431 |
| Age: Over 67 |  | -6.5 | 3.1 | [-11.5, -1.5] | [-12.5, -0.5] | 0.03375 |
| Male |  | 4.5 | 1.5 | [2.1, 7.0] | [1.7, 7.4] | 0.00201 |
| Non-binary |  | -4.7 | 5.8 | [-14.2, 4.8] | [-16.0, 6.6] | 0.41751 |
| African American or Black |  | 4.4 | 2.3 | [0.6, 8.2] | [-0.1, 9.0] | 0.05805 |
| Hispanic |  | -7.2 | 2.5 | [-11.4, -3.0] | [-12.2, -2.2] | 0.00459 |
| Asian/Pacific Islander |  | 0.4 | 3.5 | [-5.3, 6.1] | [-6.4, 7.2] | 0.90764 |
| Native American |  | -1.5 | 6.9 | [-12.8, 9.8] | [-15.0, 12.0] | 0.82793 |
| Other |  | -5.5 | 5.4 | [-14.3, 3.3] | [-16.0, 5.0] | 0.30361 |
| Multiracial |  | -6.5 | 2.8 | [-11.1, -1.8] | [-12.0, -0.9] | 0.02265 |
| College or greater |  | -0.2 | 1.9 | [-3.3, 2.9] | [-3.9, 3.5] | 0.90910 |
| Pharmacist trust index (1 SD) |  | 2.6 | 0.7 | [1.4, 3.8] | [1.2, 4.1] | < 0.001 |
| Healthcare professional |  | 3.8 | 2.9 | [-1.0, 8.6] | [-1.9, 9.6] | 0.19078 |

CI: Confidence interval; PP: percentage point; SD: standard deviation; SE: standard error
**Table note.** The outcome is an indicator for whether the respondent agreed to pay the posted price for a pharmacy service. Coefficients are from a linear probability model with service fixed effects and participant-clustered standard errors. The \$20 condition and household income are included as controls but not shown. Age categories are relative to respondents ages 18-26. Male and non-binary are each relative to female respondents. White respondents are the reference group for race/ethnicity indicators. College or greater is relative to high school diploma or below. Healthcare professional is relative to not being a healthcare professional. The pharmacist trust index is the standardized average of the nine pharmacist-trust items. Points show coefficient estimates; intervals show 90% and 95% confidence intervals.

We found several differences in payment acceptability by respondent characteristics. Older respondents were less likely to indicate that payment would be acceptable. Relative to respondents aged 18–26, respondents aged 50–67 and those older than 67 were approximately 5.4 and 6.5 percentage points less likely, respectively, to indicate that payment would be acceptable (both *p*<0.05). Estimates for respondents aged 27–34 and 35–49 were smaller and less precise.

We also observed differences by gender and race/ethnicity. Relative to female respondents, males were approximately 4.5 percentage points more likely to indicate that payment would be acceptable (*p*<0.01). Hispanic respondents were approximately 7.2 percentage points less likely to indicate that payment would be acceptable (*p*<0.01). We found marginal evidence that Black respondents were more likely to find payment acceptable (4.4 percentage points; *p*<0.10), but we did not find clear evidence of differences for Asian/Pacific Islander respondents, respondents with a college degree or greater, or respondents who identified as health care professionals.

Finally, we examined the association between trust in pharmacists and payment acceptability. A one-standard-deviation increase in the pharmacist trust index was associated with an approximately 2.6 percentage-point higher probability of indicating that payment would be acceptable (*p*<0.01). Response means for trust constructs ranged from 4.05 ± 0.86 (“makes me feel at ease”) to 4.39 ± 0.66 (“is competent”), indicating high levels of pharmacist trust across all constructs. All means for constructs of trust may be found in Supplemental Table 1.

## 5. Robustness of Agreement to Pay Results

To assess whether the service-level patterns observed in the quota-matched sample were robust to a distinct recruitment source and respondent population, we replicated the survey in an independent convenience sample of University of California San Diego (UCSD) students. The results are shown in Supplemental Figure 2. Among 218 UCSD respondents, service-level payment-acceptability rates in the $20 condition were strongly correlated with those observed in the quota-matched sample (Pearson *r* = 0.90; Spearman rho = 0.75). Service-level payment-acceptability rates in the any-payment condition were also positively correlated across samples (Pearson *r* = 0.75; Spearman rho = 0.68). Additionally, we included a $5 stated-price condition to clarify the unexpected finding that agreement was higher in the $20 stated-price condition than in the any-payment condition. Participants were more likely to indicate that the $5 price was acceptable than the $20 price, suggesting that the difference between the stated-price and any-payment conditions was driven by question wording rather than by higher prices increasing acceptability.

These results suggest that services viewed as more acceptable for direct out-of-pocket payment in the quota-matched sample also tended to be viewed as more acceptable in the UCSD sample. The replication supports the robustness of the main finding that payment acceptability varies meaningfully by service type, despite differences in respondent population and study context.

## 6. Discussion

From the outset of this study, we surmised that shifting pharmacy services from a no-charge model to an out-of-pocket payment model would have a significant impact on the market. However, an *a priori* prediction of that impact was not possible. Simple economics suggests that there would be a decrease in demand for services even if a fee should be required [14]. Alternatively, services provided for free may be viewed in the context of a “freemium” [15], a business model where some services are offered for free, but charges are imposed for more specialized services. Yet another possibility, concerns the concept of “A fine is a price,” a famous finding where adding fees to services increases rather than decreases their demand by making them more explicit [16]. In a similar vein, we wondered if imposing a fee for services by community pharmacists would result in increased patient participation.

Our survey supports the notion that a portion of the population may be willing to pay out-of-pocket for some services that could be made available at community pharmacies. This may suggest that pharmacies considering a “freemium” business model may be feasible. Under this model, if clients are accustomed to having received services at a community pharmacy for no out-of-pocket costs, evolving to payment being a required model could be considered a natural progression, and the pharmacy could build in additional goodwill. We suspect, however, the concept of “A fine is the price” is a dominant factor.

Gneezy and Rustichini essentially employed a “back door” method to ascertain an unmet need.^15^ Our survey was conducted using a “front door” method, i.e., asking the question, if health services were to be offered, would the public have the will to pay out-of-pocket costs. Overall, the current survey supports the idea that a significant portion of society is willing to consider payment for services that can be provided at a community pharmacy. The service receiving the greatest positive response, regardless of gender, was osteoporosis screening. This is important, bearing in mind recommendations that all women over the age of 65 years be screened for osteoporosis, given a prevalence of around 27% in this population. It is also recommended that postmenopausal women younger than 65 undergo screening. The benefit for men is not as robust, given a lower incidence of around 6% [17].

This is significant because the consequences of bone fragility and fracture can be severe. Throughout the world, 9.6 million hip fractures were reported in 2019, mainly in women but with increasing prevalence in men, with high one-year mortality rates (15–30%) [18]. For geriatric females following hip fracture, the following formula has been proposed for the prediction of life expectancy: (100 – Patient Age) ÷ 4 [19]. Clearly, there is significant value in identifying individuals who are at high risk and implementing a treatment plan that is directly accessible in a community setting.

It is beyond our scope to suggest independent business practices, but there are many apparent opportunities for interventions by pharmacists that could dovetail with the services suggested by our study. For example, pharmacists could improve osteoporosis management in the local community [20–21]. The FRAX® tool has been used to evaluate over 50 million individuals for fracture risk (https://www.fraxplus.org/?lang=En) [22], and measurement of calcaneal (heel) bone density using quantitative ultrasound, as can readily be performed by a pharmacist, correlates with dual-energy X-ray absorptiometry bone density [23].

Of the 27 services queried, counseling regarding antibiotic use was lowest on the list. According to the CDC, 255.9 million outpatient oral antibiotic prescriptions were dispensed in 2024 [24]. The notion of unnecessary antibiotic prescriptions [25], or overprescribing leading to antibiotic resistance [26–27], is beyond the scope of our discussion. However, given the magnitude of antibiotic distribution, it is reasonable to expect that many of those responding to this survey, as representatives of the public, have participated in some manner with antibiotic usage. In such settings, a counseling session between the patient and the pharmacist is expected and essential, even if for no other reason, to abide by the Omnibus Budget Reconciliation Act, a U.S. federal mandate that requires, among other stipulations, that a pharmacist offers to counsel patients [28]. Therefore, it is neither feasible nor ethical to dissociate dispensing from counseling, irrespective of the patient’s view on payment acceptability. Thus, the overall cost of dispensing the prescribed drug must be all-inclusive.

Between the most and least desirable services described in the survey that can be offered by community pharmacists, i.e., osteoporosis screening (highly desirable) to antibiotic counseling (less desirable), are 25 remaining services representing business opportunities that could be considered as legally allowable, with consideration that scope of practice varies by jurisdiction and that acceptability does not establish business viability. An advantage here is patient accessibility to the community pharmacist. In addition, this survey and other reports demonstrate that the community pharmacist is well respected and trusted by the public [29]. This is the first and critical step for anyone to delegate their health care to another, and our survey suggests they may be willing to do so.

The willingness and desire to receive such services represents a marketplace that will ultimately serve the public good. As described above, osteoporosis screening would clearly alleviate an unmet need. But in a similar vein, consider travel related vaccines. Perhaps 90 million or more US citizens travel to international destinations every year and based on a survey of about one million travelers reported by Lewin et al., 612,795 travelers received 898,196 doses of travel vaccines [30]. This is clearly a service that falls within the expertise of the community pharmacist, and CDC guidance of vaccination requirements is readily available as is additional training and certification by the American Pharmacists Association [31].

Another notable example is assisting with diabetic foot care. Over 10% of the US population is affected by diabetes, around 15% of those will develop foot ulcers, and around 20% of those will require amputation [32]. Here, the community pharmacist can have an impact on quality of life and health outcomes. On the front line, the community pharmacist has direct access to patients with diabetes. Methods of counseling and promoting foot care health are well within the scope of practice [33–34]. Our survey indicates that patients would be willing to pay for this service, and it is reasonable to expect that such counseling would lead to improved health and wellbeing.

Naloxone counseling and distribution is yet another clinical service that could be easily rendered in the community pharmacy. With opioid-related deaths of great concern across the United States, pharmacists are well-positioned to identify potential at-risk patients and caregivers who would benefit from training on the administration of naloxone while destigmatizing the issue and the opioid crisis. According to National Association of Boards of Pharmacy (NABP), pharmacists have become an essential part of the country’s strategy to combat the opioid epidemic [35]. Naloxone distribution is a key factor in contributing to a decline in opioid overdose deaths. Pharmacists remain well positioned to continue outreach efforts, education, and distribution in the pharmacy and the community. This service, having an impact beyond the walls of the community pharmacy, is an investment many respondents were willing to pay.

Administration of long-acting injectables (LAI), such as extended-release naltrexone or antipsychotics, was a clinical service that ranked high in fee acceptability. Although this service has been in practice for some time, not all pharmacies offer it due to logistics, increase in training, and variable patient demand. Mooney *et al*. evaluated patient satisfaction with community pharmacist administered LAI antipsychotics and found patients were satisfied and found it more convenient than other settings [36]. Advantages of LAI administration in the community pharmacy span across many domains including increased access and adherence, improved continuity of care, medication therapy management, decreased stigma and increased acceptance in a familiar setting, and early intervention for missed doses. Overall, patient outcomes are improved with increased access and adherence. Bustamante *et al.* reported the need to expand patient access to LAI for substance use disorder in the pharmacy setting as there is scant literature on this clinical service in the community pharmacy [37].

Point-of-care testing (POCT) is another area in which respondents were willing to pay a fee for in the community pharmacy. POCT involves performing rapid diagnostic tests in a convenient setting close to patient care sites without sending samples to an outside lab and may encompass tests for COVID-19, influenza A/B, streptococcus A, and RSV with treatment protocols in many cases. Monitoring for glycosolated hemoglobin (A1c), cholesterol levels, and INRs are other POCT that can be performed in the community pharmacy, depending on the state. Increased access to services, convenience, cost, and patient acceptance improves health outcomes for patients and communities. This service may expand in the future as POCT grows into the pharmacogenomics domain and becomes available for use and interpretation in the community pharmacy [38]. Increased education and training will need to coincide with this expansion, but community pharmacists are well positioned to meet the needs of increased POCT. Additional literature suggests that worldwide, pharmacists face similar challenges in providing services without remuneration [39].

Strengths of the current survey include its large quota-matched sample size. In the U.S., there are approximately 270 million adults over the age of 18. To perform a survey with a desirable 95% confidence level and 3% margin of error, a standard sample size formula produces the requirement of 1067 participants. This is the number of participants we selected for this survey. Naturally, however, to be representative of the entire population, it would be necessary to randomly sample based on proper proportions of individuals who reasonably represent the entire U.S. adult population, particularly regarding age, gender, race and ethnicity, education, income, and geographic distribution. This is not a simple task but, given the relatively large number of survey takers (>300,000) associated with the Prolific instrument, we assumed our resulting data would be adequate to answer the key theories of the work, perhaps with some qualifications.

First, the educational level of survey respondents differed from that of the general U.S. adult population. Approximately 81.8% of survey respondents reported at least some college education (table 2), compared with approximately 62.7% of U.S. adults aged 18 years and older [40]. Conversely, 18.2% of survey respondents reported a high school education or less, compared with approximately 37.3% of the U.S. adult population. Future research should examine whether educational attainment influences perceptions of pharmacist-provided services and stated payment acceptability. Nonetheless, our survey included representation of a large swath of the U.S.

Another variable that may influence the willingness and ability to pay directly for community pharmacist-provided services is household income. As shown in Supplemental Table 2, in our survey, the proportion of respondents reporting annual household incomes below $30,000, and $75,000–$149,999, closely aligned with national estimates [41]. However, respondents reporting household incomes between $30,000 and $149,999 were overrepresented, whereas those reporting incomes of $150,000 or more were underrepresented. Consequently, when evaluating the generalizability of our findings to the broader U.S. population, the income distribution of our sample needs to be considered.

An especially strong feature of our survey is the geographic representation of the respondents [42]. As shown in Supplemental Table 3, only small differences between our sample versus the population distribution were discerned. Accordingly, we conclude that our sample achieves good geographic coverage.

Similarly, our survey participants show excellent alignment with the US population in terms of sex (male and female) and younger age groups (ages 18–26, 27–34, and 35–49) (see Supplemental Table 4) [43]. Also, the total percentage of respondents in the age group of 50 to over 65 (46%) aligns perfectly with the general population in this age range (45%). However, examination of the more narrowly defined age groups of 50-65 and over 65 shows over representation of 7% and underrepresentation of 6%, respectively. Interestingly, relative to the 18-26 age group, respondents aged 50–67 and those older than 67 were approximately 5.4 and 6.5 percentage points less likely to indicate payment would be acceptable. These percentages are not of sufficient magnitude to change the overall interpretation of this study, but further refinement would be of interest.

Lastly, the percentage of participants identifying as Blacks and Asians closely align with the US population (Supplemental Table 4). Ostensibly, however, Hispanic respondents were underrepresented given that 8% self-identified in this group whereas the Census specifies 17%. The point is somewhat moot given that our survey treats Hispanic identity as a race category, whereas the Census measures Hispanic origin separately from race. Nonetheless, in our survey, those identifying as Hispanic respondents were approximately 7.2 percentage points less likely to indicate that payment would be acceptable (p<0.01). If interest in this point persists, the matter would not be difficult to resolve in a future survey.

Finally, some potential limitations of our study may be noted. First, although Prolific touts a large number of survey participants (approximately 300,000), recruiting participants through this mechanism rather than random sampling may limit the generalizability of our findings. Second, although we considered asking participants if they themselves would pay for the respective services, knowing ahead of time that many of them personally would not require the respective services hypothetical bias may affect responses, as participants might answer differently when faced with actual payment decisions. Next, although the responses were revealing, the framing of the any-payment condition may have introduced ambiguity, as participants could interpret “any amount” as either some positive payment or an unrestricted amount, which was not the intention. Lastly, even though our survey correlated very well with other surveys that have been designed to assess trust in community pharmacists, since trust was measured after participants evaluated the 27 services, this exposure may have influenced their trust ratings and the observed relationship between trust and payment acceptability.

## 7. Conclusion

The pharmacist is a highly skilled healthcare professional who plays an integral role in the U.S. healthcare system. Our goal was to query services within the entire potential scope of U.S. pharmacist licensure that would meet the needs of the public and could be offered in the immediate future based on the will of the pharmacist and current business models We surmise that any major shift in the current paradigm of community pharmacy services would likely be influenced by the will of the people and the ability to receive payment for service. Our survey results strongly support the notion that a significant portion of the general population believes that people in need of specific services that could conveniently be provided at a community pharmacy would find such services to be of sufficient value to pay out-of-pocket. Naturally, additional surveys can be constructed and administered to acquire additional evidence to support or dispute this payment model. In our view, however, the next best step would be acquisition of definitive evidence through implementation of pilot projects in select market areas where public health improvements could be realized.

## Supporting information

Supplemental for Pesaturo et al.

## Data Availability

All data produced in the present work are contained in the manuscript

^1^We note that the 1922 code was in effect until 1952, and the current code APhA Code of Ethics for Pharmacists can be found here: https://www.pharmacist.com/Code-of-Ethics.

## Declarations

## Funding

This research was funded internally. The research did not receive any specific grant from funding agencies in the public, commercial, or not-for-profit sectors.

## Conflict of interest

The authors have no relevant financial or non-financial interests to disclose.

## Ethics approval

This study was reviewed by the Institutional Review Board of Western New England University (No. 316) and the University of California, San Diego (No. 812140). The studies were performed in accordance with the Declaration of Helsinki (1964) and its subsequent amendments.

## Consent for publication

Informed consent was obtained from all individual participants included in the study. Participants under age 18 were not able to participate in this study.

## Availability of data and material (data transparency)

Pezzuto, John-Henry. 2026. “ Replication Package for: Public Acceptability of Self-Payment for Community Pharmacists’ Services” Harvard Dataverse. https://doi.org/10.7910/DVN/ZFAVPQ.

## Code availability

Pezzuto, John-Henry. 2026. “Replication Package for: Public Acceptability of Self-Payment for Community Pharmacists’ Services.” Harvard Dataverse. https://doi.org/10.7910/DVN/ZFAVPQ.

## Authors’ contribution

All authors read and approved the final version. **Kimberly A. Pesaturo**: conceptualization, methodology, validation, investigation, formal analysis, writing – original and revised draft, writing – review and editing; **John-Henry Pezzuto**: conceptualization, methodology, validation, formal analysis, writing – original and revised draft, writing – review and editing, visualization; **Melissa Mattison**: conceptualization, methodology, writing – original and revised draft, writing – review and editing; **John Pezzuto**: conceptualization, methodology, investigation, writing – original and revised draft, writing – review and editing, supervision, funding acquisition

## AI use

Generative AI tools were used to assist with manuscript editing, code development, figure preparation, and comparisons between the survey sample and the U.S. population. The authors reviewed the AI-assisted outputs and verified population statistics against official U.S. Census sources. The authors take full responsibility for the final manuscript.

## Notes

### Competing Interest Statement

The authors have declared no competing interest.

### Author Declarations

The Institutional Review Board of Western New England University reviewed with study (No. 316) and ethical approval was given. The Institutional Review Board of the University of California, San Diego reviewed this study (No. 812140) and approval was given.

