## Supplemental for Pesaturo et al. for "Public Acceptability of Self-Payment for Community Pharmacists’ Services: A National Online Survey"

**Supplement Fig.1.** Rate agreeing to pay posted price by service, condition, and gender.

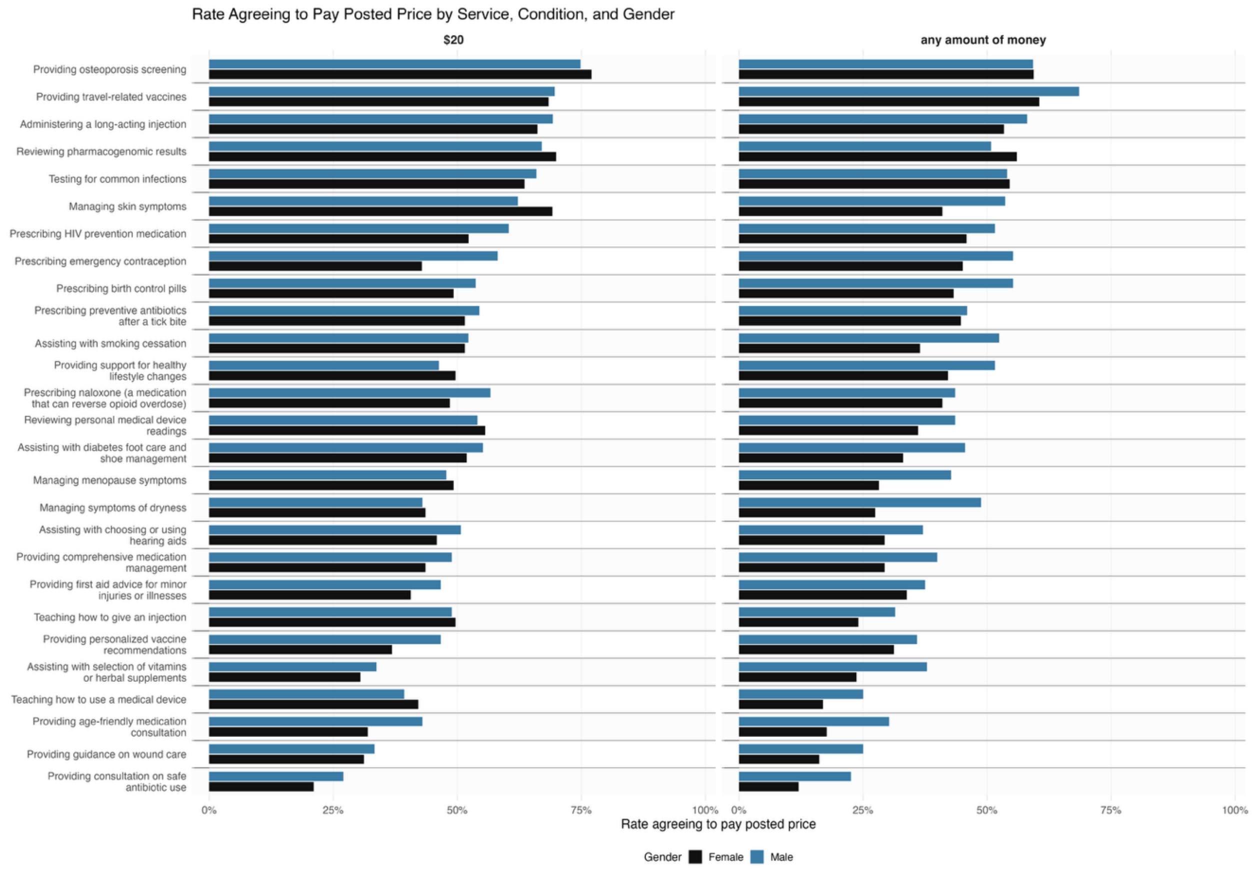


**Figure note.** Participants evaluated 27 pharmacist-provided services and indicated whether they agreed to pay the posted out-of-pocket price for each service. Participants in the main sample were randomly assigned to one of two payment conditions: $20 out-of-pocket payment or any out-of-pocket payment. The figure shows the proportion agreeing to pay within each service, condition, and gender subgroup. The gender-stratified analysis includes respondents identifying as female or male: $20 condition, 266 female and 270 male respondents; any out-of-pocket payment condition, 266 female and 248 male respondents.

**Supplement Fig.2.** Rate agreeing to pay posted price by condition, UCSD Sample.

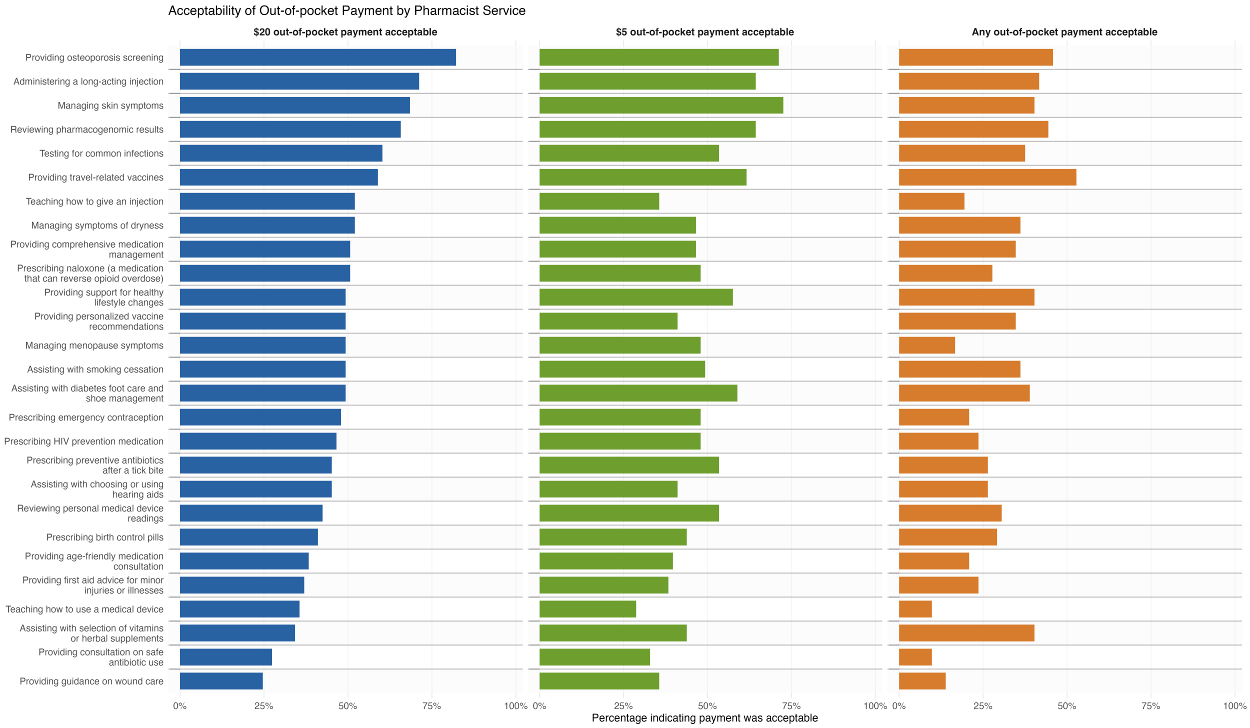


**Figure note.** University of California San Diego (UCSD) student participants evaluated 27 pharmacist-provided services and indicated whether the posted out-of-pocket payment amount was acceptable for each service. Participants were randomly assigned to one of three payment conditions: $20 out-of-pocket payment, $5 out-of-pocket payment, or any out-of-pocket payment. The analytic sample included 218 students: 73 assigned to the $20 condition, 73 assigned to the $5 condition, and 72 assigned to the any out-of-pocket payment condition. Students participated in the study for course credit.

**Supplement Table 1**. Mean agreement among constructs of trust.

| **Construct (n responses, response %)[a]**<br><br>**"The pharmacist…** | **Response (mean ± std dev)[b]** |
|---|---|
| provides kindness/positive intention." (1014, 94.8) | 4.14 ± 0.81 |
| is competent." (1000, 93.5) | 4.39 ± 0.66 |
| works with integrity." (984, 92.0) | 4.29 ± 0.71 |
| makes me feel at ease." (1002, 93.7) | 4.05 ± 0.86 |
| is fair." (977, 91.4) | 4.23 ± 0.72 |
| communicates well." (988, 92.4) | 4.24 ± 0.78 |
| is dependable." (1001, 93.6) | 4.27 ± 0.78 |
| is confidential." (967, 90.5) | 4.35 ± 0.72 |
| I trust the pharmacist. (995, 93.1) | 4.26 ± 0.76 |

[a]n responses excludes "no basis for observation"

[b]see Supplement Figure 1 for definition or the Likert scale options 1-5.

std dev: standard deviation

**Supplement Figure 3. Prolific Survey**

**Supplement Figure 4.  Replication Study Survey**

### Part 2: full survey design

### Part 2: the full survey with all variations, including validation, randomization rules, js code, etc.

**Text styles used below**

| | |
|---|---|
| ‣ *Green text* | *Rules participants do not see: display logic, branches, randomization, validation, timers.* |
| `text1` | `Text dynamically generated during the survey by randomization or past responses.` |
| red 1A<br>blue 1B | Text that differs by randomly assigned condition. The small label names the condition (1A, 1B). |

‣ *Back button: not shown.*

‣ *At the end, participants are redirected to https://app.prolific.com/submissions/complete?cc=C2VNGQYR.*

6 blocks · 36 questions · 1 randomizer

‣ Random assignment #1: each participant sees one of the following 2 conditions (evenly presented).

1A <low> = 1
<X1> = "any amount of money"

1B <low> = 0
<X1> = "$20"

*End of random assignment #1*

**Block 1** Consent

-- Screen #38 --

QConsent

Title: Assessment of Public Perception of Trust and Reimbursement for Community Pharmacist Services

Primary Investigator: Kimberly Pesaturo

Secondary Investigators: John-Henry Pezzuto, Melissa Mattison, John Pezzuto

The goal of this research is to identify perceptions surrounding trust for payment of services provided by community pharmacists. You are invited to participate in this research project because you have been identified as a qualified survey participant through the parent company, Prolific, or through on-site recruitment.

Participation in this research study is voluntary, and the survey will be provided electronically after you agree to this consent. You may choose not to participate. Participation is anonymous, and there is minimal risk involved. You may change your mind and withdraw from the study at any time before submitting the survey without any penalty. If you decide not to participate or withdraw, your decision will have no impact on any future actions with or from the investigators.

The procedure involves completing an online survey. The survey should take approximately 10 minutes to complete. You will be asked 27 brief questions about potential payment for individual services that could be provided in the community pharmacy. Additionally, you will be asked about perceptions of trust regarding these services and some brief demographics questions.

All survey responses will be pooled, and no identifiable data will be collected. All data will be stored securely in a password-protected file for six years, accessible only to the research team. The results of this study may be used for scholarly purposes, such as presentations or publications.

By selecting "Agree," you confirm the following:
- You have read and understood the above information.
- You voluntarily agree to participate.
- You are at least 18 years of age.
- You are able to answer the survey in English.
- You are based in the United States.
- You are not a prisoner.

If you have any questions about the research study, please contact Dr. Kimberly Pesaturo, the Principal Investigator, at or 413-796-2473. This study has been approved by the Western New England University Institution Review Board (IRB) through the College of Pharmacy and Health Sciences. If you have questions about your rights as a research participant, please contact Susan Agostine, College of Pharmacy IRB, at, or Jessica Carlson, IRB Chair, at.

By choosing "I consent," you agree to take part in the study and agree to the terms outlined in the informed consent.

○ I consent.

○ I do not consent.

▸ *Response required.*

▸ *Note 1: if the participant answers {I do not consent.}, the survey ends (later questions are not shown).*

---

▸ *Hidden question (QID140) records browser, operating system and screen resolution.*

→

If <PROLIFIC_PID> is empty then:

**Block 2** Prolific Validation

-- Screen #39 --

Q213

To see if you are eligible to participate, please enter your Prolific ID into the box below and then click NEXT.

Your Prolific ID contains letters and numbers and is 24 characters long. It has no spaces. It is NOT your email address.

*Response required.*

*Validation: total char.*

→

*End of condition*

**Block 3** Overview

-- Screen #40 --

Q138

The questions in this survey you will answer 27 main questions about services provided by pharmacies. You will then be asked to complete a short follow-up survey.

For completing this study, you will receive approximately $1.80 anticipated within 24 hours. Payment will be provided only for surveys completed in full.

**Please answer all questions carefully and honestly.**

Q196

**Understanding Question**: How should you answer the 27 main questions?

- Randomly
- Carefully and honestly
- It doesn't matter

*Response required.*

*Custom validation: answer accepted only if* <Q196> *("Understanding Question : How should you answer the ${e://Field/n_services} main questions?")* = "Carefully and honestly".

*Hidden page timer (Q133) records time spent on this page.*

→

-- Screen #41 --

Q198

That's correct!

Please continue.

→

-- Screen #42 --

Q209

Please check the box to verify that you are not a robot.

☐ I'm not a robot reCAPTCHA

→

**Block 4** WTP services instructions

-- Screen #43 --

Q195

**Instructions**

In this survey, you will evaluate 27 **health services** that may be provided by a community pharmacy.

For each health service, please indicate if it would be acceptable for a typical patient to pay at the selected price**.**

"Out-of-pocket" refers to the amount paid **directly by the patient** (such as a deductible or co-payment), not covered by insurance.

Q199

**Understanding Question**: When answering the following questions, who is paying the out-of-pocket amount?

- ◯ The typical patient
- ◯ Me
- ◯ The insurance company
- ◯ The pharmacy

*Response required.*

*Custom validation: answer accepted only if* <Q199> *("Understanding Question : When answering the following questions, who is paying the out-of-pocket amount?")* = "The typical patient".

Q204

**Understanding Question**: What does out-of-pocket refer to?

- ◯ The amount paid directly by the patient (such as a deductible or co-payment)
- ◯ The total amount the pharmacist charges
- ◯ The amount covered by insurance
- ◯ The amount paid by the insurance company

*Response required.*

*Custom validation: answer accepted only if* <Q204> *("Understanding Question : What does out-of-pocket refer to?")* = "The amount paid directly by the patient (such as a deductible or co-payment)".

→

-- Screen #44 --

Q202

That's correct!

Please continue.

→

**Block 5** Willingness to pay

*The same 3 questions are asked of each item in this loop.*

Loop & merge: this block repeats 27 times in random order. It is shown below for the first loop; highlighted loop fields take these values in each repetition (1 loop field(s) not used in this block are omitted):

| # | Field 2 | Field 3 |
|---|---|---|
| 1 | Teaching how to use a medical device | Teaches patients how to properly use a medical device (such as an injection, inhaler, blood sugar monitor, or blood pressure cuff) in a single consultation. |
| 2 | Teaching how to give an injection | Teaches patients how to safely and correctly give themselves a prescribed injection (such as insulin or epinephrine) in a single consultation. |
| 3 | Reviewing personal medical device readings | Reviews patients' medical device readings (such as blood sugar readings or blood pressure values) and provides guidance on managing their associated condition in a single consultation. |
| 4 | Providing support for healthy lifestyle changes | Provides guidance to improve patients' diet, exercise, or weight management in a single consultation. |
| 5 | Providing comprehensive medication management | Reviews all of patients' medications to improve safety and effectiveness and recommends modifications as needed. |
| 6 | Assisting with choosing or using hearing aids | Assists patients with selecting and properly using their hearing aid in a single consultation. |
| 7 | Managing symptoms of dryness | Evaluates and provides treatment recommendations for patients' symptoms of dryness (such as dry eyes or dry mouth) in a single consultation. |
| 8 | Managing skin symptoms | Evaluates and provides treatment recommendations for patients' skin symptoms (such as rash, eczema, or acne) in a single consultation. |
| 9 | Providing first aid advice for minor injuries or illnesses | Provides advice and treatment recommendations for patients' minor injuries or illnesses in a single consultation. |
| 10 | Providing personalized vaccine recommendations | Reviews patients' vaccination history, recommends, and provides necessary vaccines in a single consultation. The service fee is charged separately from the cost of the vaccines. |
| 11 | Providing travel-related vaccines | Recommends and administers vaccines needed for patients' travel destinations in a single consultation. The service fee is charged separately from the cost of the vaccines. |
| 12 | Managing menopause symptoms | Evaluates and provides treatment recommendations for patients' menopause symptoms in a single consultation. |
| 13 | Providing osteoporosis screening | Conducts one-time screening to assess patients' bone strength and risk of osteoporosis. |
| 14 | Providing consultation on safe antibiotic use | Reviews patients' prescribed antibiotics for appropriateness and explains how to use them safely in a single consultation. |
| 15 | Providing age-friendly medication consultation | Provides a one-time consultation of medications to ensure they are safe and appropriate based on patients' age. |
| 16 | Prescribing naloxone (a medication that can reverse opioid overdose) | Prescribes naloxone one time and explains how patients can use it in case of an opioid overdose. The service fee is charged separately from the cost of naloxone. |
| 17 | Prescribing HIV prevention medication | Prescribes a supply of medication that helps prevent HIV infection and explains proper use. The service fee is charged separately from the cost of the medication. |

| # | Field 2 | Field 3 |
|---|---|---|
| 18 | Prescribing birth control pills | Prescribes a supply of birth control pills and explains how patients should use them safely. The service fee is charged separately from the cost of the birth control pills. |
| 19 | Prescribing emergency contraception | Prescribes one-time emergency contraception and explains when and how it should be taken. The service fee is charged separately from the cost of the emergency contraception. |
| 20 | Prescribing preventive antibiotics after a tick bite | Prescribes preventive antibiotics and explains how to take them after a tick bite consultation. The service fee is charged separately from the cost of the antibiotics. |
| 21 | Testing for common infections | Tests patients for infections such as strep throat, COVID, or flu based on symptoms and explains the results. The service fee is charged separately from the cost of the test. |
| 22 | Assisting with selection of vitamins or herbal supplements | Advises patients on choosing safe and appropriate vitamins or herbal supplements in a single consultation. |
| 23 | Administering a long-acting injection | Provides a one-time injection and follow-up care for patients on long-acting injectable medication. The service fee is charged separately from the cost of the medication. |
| 24 | Assisting with diabetes foot care and shoe management | Assists with product selection for general foot care, therapeutic shoes, and/or inserts in a single consultation. |
| 25 | Reviewing pharmacogenomic results | Evaluates DNA testing kit results to provide personalized advice related to medications in a single consultation. |
| 26 | Assisting with smoking cessation | Assists patients with product selection and smoking cessation management in a single consultation. |
| 27 | Providing guidance on wound care | Advises patients on appropriate product selection for wound care in a single consultation. |

-- Screen #45 --

QID143

**Service [loop field CurrentLoopNumber] out of [loop field TotalLoops]:**

The following service may be provided by a community pharmacy:

`Teaching how to use a medical device`

For this service the pharmacist:

`Teaches patients how to properly use a medical device (such as an injection, inhaler, blood sugar monitor, or blood pressure cuff) in a single consultation.`

wtp1_raw

**In your view, for a typical patient, would it be acceptable for the patient to pay** `any amount of money` [1A]
`$20` [1B] **out-of-pocket for this service?**

- Yes
- No

*Response required.*

*JavaScript records embedded data fields: agree1_, order_.*

*Hidden page timer (Q135) records time spent on this page.*

→

**Block 6** Follow up survey

-- Screen #46 --

Q200

**To finish the study, please complete the follow-up survey questions on the next three pages.**

**Please answer these questions carefully and honestly.**

→

-- Screen #47 --

QID145

**Follow-up survey (Page 1 out of 3)**

Q1

Please rate your agreement with the following statements about your local community pharmacist:

| | Strongly Disagree | Disagree | Neutral | Agree | Strongly Agree | No basis for observation |
|---|---|---|---|---|---|---|
| The pharmacist provides kindness/positive intention. | ◯ | ◯ | ◯ | ◯ | ◯ | ◯ |
| The pharmacist is competent. | ◯ | ◯ | ◯ | ◯ | ◯ | ◯ |
| The pharmacist works with integrity. | ◯ | ◯ | ◯ | ◯ | ◯ | ◯ |
| The pharmacist makes me feel at ease. | ◯ | ◯ | ◯ | ◯ | ◯ | ◯ |
| The pharmacist is fair. | ◯ | ◯ | ◯ | ◯ | ◯ | ◯ |
| The pharmacist communicates well | ◯ | ◯ | ◯ | ◯ | ◯ | ◯ |
| The pharmacist is dependable. | ◯ | ◯ | ◯ | ◯ | ◯ | ◯ |
| The pharmacist is confidential. | ◯ | ◯ | ◯ | ◯ | ◯ | ◯ |
| I trust the pharmacist | ◯ | ◯ | ◯ | ◯ | ◯ | ◯ |

▸ *Response required.*

▸ *Hidden page timer (Q164) records time spent on this page.*

→

-- Screen #48 --

Q165

**Follow-up survey (Page 2 out of 3)**

38.

What is the approximate distance from your home to the nearest pharmacy? (miles)

- Less than 1
- 2 - 5
- 6 - 10
- Greater than 10

*Response required.*

39.

When was your last visit to a pharmacy?

- Within 3 months
- 3 - 6 months
- Greater than 6 months

*Response required.*

40.

Do you have a diagnosed chronic medical condition (defined here as a condition requiring medication to be routinely refilled)?

- Yes
- No

*Response required.*

*Hidden page timer (Q166) records time spent on this page.*

→

-- Screen #49 --

Q201

**Follow-up survey (Page 3 out of 3)**

41.

Please indicate how much you agree with the following statements.

| | Strongly Disagree | Disagree | Neutral | Agree | Strongly Agree |
|---|---|---|---|---|---|
| I made each decision in this study carefully. | ◯ | ◯ | ◯ | ◯ | ◯ |
| I made decisions in this study randomly. | ◯ | ◯ | ◯ | ◯ | ◯ |
| I understood what my decisions meant for how much a patient may pay. | ◯ | ◯ | ◯ | ◯ | ◯ |
| Select the button that is furthest to the left. | ◯ | ◯ | ◯ | ◯ | ◯ |
| Select the button that is furthest to the right. | ◯ | ◯ | ◯ | ◯ | ◯ |

▸ *Response required.*

healthcare

Do you consider yourself a healthcare professional?

◯ Yes

◯ No

▸ *Response required.*

age

How old are you? (years)

◯ 18 - 26

◯ 27 - 34

◯ 35 - 49

◯ 50 - 67

◯ Over 67

▸ *Response required.*

gender

What is your stated gender?

◯ Male

◯ Female

◯ Non-binary

▸ *Response required.*

race

What is your stated race? (Select all that apply.)

- ☐ Caucasian or white
- ☐ Hispanic
- ☐ African American or Black
- ☐ Asian or Pacific Islander
- ☐ Native American
- ☐ Other

*Response required.*

marriage

What is your current relationship status?

- ◯ Single
- ◯ Married
- ◯ Other (e.g., partnered or dating, separated, divorced, widowed)

*Response required.*

education

What is your highest level of formal education?

- ◯ High school diploma or below
- ◯ College diploma or some college
- ◯ Post-graduate

*Response required.*

income

What is your annual pre-tax household income?

- ◯ Less than $30,000
- ◯ $30,000 - 74,999
- ◯ $75,000 - 149,999
- ◯ More than $150,000

*Response required.*

region

What is your primary geographic location?

- ◯ Northeast - New England (CT, ME, MA, NH, RI, VT)
- ◯ Northeast - Mid-Atlantic (NJ, NY, PA)
- ◯ Midwest - East North Central (IL, IN, MI, OH, WI)
- ◯ Midwest - West North Central (IA, KS, MN, MO, NE, ND, SD)
- ◯ South - South Atlantic (DE, FL, GLA, MD, NC, SC, VA, WV)
- ◯ South - East South Central (AL, KY, MS, TN)
- ◯ South - West South Central (AR, LA, OK, TX)
- ◯ West - Mountain (AZ, CO, ID, MT, NV, NM, UT, WY)
- ◯ West - Pacific (AK, CA, HI, OR, WA)

▸ *Response required.*

option_text

(Optional): If you would like to, please describe how you made decisions in this study.

▸ *Hidden page timer (Q205) records time spent on this page.*

→

▸ End of survey: participants are redirected to https://app.prolific.com/submissions/complete?cc=C2VNGQYR.

### Part 2: the full survey with all variations, including validation, randomization rules, js code, etc.

**Text styles used below**

| | |
|---|---|
| ▸ *Green text* | *Rules participants do not see: display logic, branches, randomization, validation, timers.* |
| `text1` | `Text dynamically generated during the survey by randomization or past responses.` |
| red 1A<br>blue 1B | Text that differs by randomly assigned condition. The small label names the condition (1A, 1B). |

▸ *Back button: not shown.*

▸ *At the end, participants are redirected to https://ucsd.co1.qualtrics.com/jfe/form/SV_6zXKN99kyr14PEa.*

6 blocks · 31 questions · 1 randomizer

▸ Random assignment #1: each participant sees one of the following 3 conditions (evenly presented).

1A <low> = 1
<X1> = "any amount of money"

1B <low> = 1
<X1> = "$5"

1C <low> = 0
<X1> = "$20"

*End of random assignment #1*

**Block 1** SONA ID

-- Screen #39 --

Q216

Please continue with the following survey, this is **NOT** the end of the lab:

Q213

Please enter your 5 or 6-digit SONA ID:

▸ *Response required.*

▸ *Answer length must be at least 5 characters.*

→

**Block 2** Consent

-- Screen #40 --

QConsent

You are being asked to take part in a research study.

The purpose of this research is to better understand decision-making processes.

Your participation in this study is voluntary and you may withdraw your participation at any time for any reason.

If you take part in this study, you will be asked to answer several questions and answer a short follow-up questionnaire. Your participation will take approximately `10` minutes to complete. You will receive clear instructions and will be provided with full and accurate information on how your answers will impact your payment. You may not be told everything about the purpose of the study. None of the data you provide in this study will be linked to any information that could identify you to others. In the future, this non-identifiable data may be shared with other researchers or participants.

We do not anticipate any serious risks or discomfort from participating in this study.

We cannot promise any benefits to you or others from your taking part in this research. However, possible benefits include the importance of knowledge to be gained for society at large.

You can decline to participate in any part of this study for any reason and can end your participation at any time.

If you have any questions about this study, you can contact John-Henry Pezzuto at.

Thank you again for your time and participation.

If you would like to voluntarily participate in this survey and confirm that you are 18 years or older, please press the arrow to continue.

▸ *Hidden question (QID140) records browser, operating system and screen resolution.*

→

**Block 3** Overview

-- Screen #41 --

Q138

The questions in this survey you will answer 27 main questions about services provided by pharmacies. You will then be asked to complete a short follow-up survey.

**Please answer all questions carefully and honestly.**

Q196

**Understanding Question**: How should you answer the 27 main questions?

- ◯ Randomly
- ◯ Carefully and honestly
- ◯ It doesn't matter

*Response required.*

*Custom validation: answer accepted only if* <Q196> *("Understanding Question : How should you answer the ${e://Field/n_services} main questions?")* = "Carefully and honestly".

*Hidden page timer (Q133) records time spent on this page.*

→

-- Screen #42 --

Q198

That's correct!

Please continue.

→

-- Screen #43 --

Q209

Please check the box to verify that you are not a robot.

☐ I'm not a robot reCAPTCHA

→

**Block 4** WTP services instructions

-- Screen #44 --

Q195

#### Instructions

In this survey, you will evaluate 27 **health services** that may be provided by a community pharmacy.

For each health service, please indicate if it would be acceptable for a typical patient to pay at the selected price**.**

"Out-of-pocket" refers to the amount paid **directly by the patient** (such as a deductible or co-payment), not covered by insurance.

---

Q199

**Understanding Question**: When answering the following questions, who is paying the out-of-pocket amount?

- ◯ The typical patient
- ◯ Me
- ◯ The insurance company
- ◯ The pharmacy

▸ *Response required.*

▸ *Custom validation: answer accepted only if* <Q199> *("Understanding Question : When answering the following questions, who is paying the out-of-pocket amount?")* = "The typical patient".

---

Q204

**Understanding Question**: What does out-of-pocket refer to?

- ◯ The amount paid directly by the patient (such as a deductible or co-payment)
- ◯ The total amount the pharmacist charges
- ◯ The amount covered by insurance
- ◯ The amount paid by the insurance company

▸ *Response required.*

▸ *Custom validation: answer accepted only if* <Q204> *("Understanding Question : What does out-of-pocket refer to?")* = "The amount paid directly by the patient (such as a deductible or co-payment)".

→

-- Screen #45 --

Q202

That's correct!

Please continue.

→

**Block 5** Willingness to pay

*The same 3 questions are asked of each item in this loop.*

Loop & merge: this block repeats 27 times in random order. It is shown below for the first loop; highlighted loop fields take these values in each repetition (1 loop field(s) not used in this block are omitted):

| # | Field 2 | Field 3 |
|---|---|---|
| 1 | Teaching how to use a medical device | Teaches patients how to properly use a medical device (such as an injection, inhaler, blood sugar monitor, or blood pressure cuff) in a single consultation. |
| 2 | Teaching how to give an injection | Teaches patients how to safely and correctly give themselves a prescribed injection (such as insulin or epinephrine) in a single consultation. |
| 3 | Reviewing personal medical device readings | Reviews patients' medical device readings (such as blood sugar readings or blood pressure values) and provides guidance on managing their associated condition in a single consultation. |
| 4 | Providing support for healthy lifestyle changes | Provides guidance to improve patients' diet, exercise, or weight management in a single consultation. |
| 5 | Providing comprehensive medication management | Reviews all of patients' medications to improve safety and effectiveness and recommends modifications as needed. |
| 6 | Assisting with choosing or using hearing aids | Assists patients with selecting and properly using their hearing aid in a single consultation. |
| 7 | Managing symptoms of dryness | Evaluates and provides treatment recommendations for patients' symptoms of dryness (such as dry eyes or dry mouth) in a single consultation. |
| 8 | Managing skin symptoms | Evaluates and provides treatment recommendations for patients' skin symptoms (such as rash, eczema, or acne) in a single consultation. |
| 9 | Providing first aid advice for minor injuries or illnesses | Provides advice and treatment recommendations for patients' minor injuries or illnesses in a single consultation. |
| 10 | Providing personalized vaccine recommendations | Reviews patients' vaccination history, recommends, and provides necessary vaccines in a single consultation. The service fee is charged separately from the cost of the vaccines. |
| 11 | Providing travel-related vaccines | Recommends and administers vaccines needed for patients' travel destinations in a single consultation. The service fee is charged separately from the cost of the vaccines. |
| 12 | Managing menopause symptoms | Evaluates and provides treatment recommendations for patients' menopause symptoms in a single consultation. |
| 13 | Providing osteoporosis screening | Conducts one-time screening to assess patients' bone strength and risk of osteoporosis. |
| 14 | Providing consultation on safe antibiotic use | Reviews patients' prescribed antibiotics for appropriateness and explains how to use them safely in a single consultation. |
| 15 | Providing age-friendly medication consultation | Provides a one-time consultation of medications to ensure they are safe and appropriate based on patients' age. |
| 16 | Prescribing naloxone (a medication that can reverse opioid overdose) | Prescribes naloxone one time and explains how patients can use it in case of an opioid overdose. The service fee is charged separately from the cost of naloxone. |
| 17 | Prescribing HIV prevention medication | Prescribes a supply of medication that helps prevent HIV infection and explains proper use. The service fee is charged separately from the cost of the medication. |

| # | Field 2 | Field 3 |
|---|---|---|
| 18 | Prescribing birth control pills | Prescribes a supply of birth control pills and explains how patients should use them safely. The service fee is charged separately from the cost of the birth control pills. |
| 19 | Prescribing emergency contraception | Prescribes one-time emergency contraception and explains when and how it should be taken. The service fee is charged separately from the cost of the emergency contraception. |
| 20 | Prescribing preventive antibiotics after a tick bite | Prescribes preventive antibiotics and explains how to take them after a tick bite consultation. The service fee is charged separately from the cost of the antibiotics. |
| 21 | Testing for common infections | Tests patients for infections such as strep throat, COVID, or flu based on symptoms and explains the results. The service fee is charged separately from the cost of the test. |
| 22 | Assisting with selection of vitamins or herbal supplements | Advises patients on choosing safe and appropriate vitamins or herbal supplements in a single consultation. |
| 23 | Administering a long-acting injection | Provides a one-time injection and follow-up care for patients on long-acting injectable medication. The service fee is charged separately from the cost of the medication. |
| 24 | Assisting with diabetes foot care and shoe management | Assists with product selection for general foot care, therapeutic shoes, and/or inserts in a single consultation. |
| 25 | Reviewing pharmacogenomic results | Evaluates DNA testing kit results to provide personalized advice related to medications in a single consultation. |
| 26 | Assisting with smoking cessation | Assists patients with product selection and smoking cessation management in a single consultation. |
| 27 | Providing guidance on wound care | Advises patients on appropriate product selection for wound care in a single consultation. |

-- Screen #46 --

QID143

**Service [loop field CurrentLoopNumber] out of [loop field TotalLoops]:**

The following service may be provided by a community pharmacy:

`Teaching how to use a medical device`

For this service the pharmacist:

`Teaches patients how to properly use a medical device (such as an injection, inhaler, blood sugar monitor, or blood pressure cuff) in a single consultation.`

wtp1_raw

**In your view, for a typical patient, would it be acceptable for the patient to pay** any amount of money [1A]
$5 [1B]
$20 [1C] **out-of-pocket for this service?**

- Yes
- No

*Response required.*

*JavaScript records embedded data fields: agree1_, order_.*

*Hidden page timer (Q135) records time spent on this page.*

→

**Block 6** Follow up survey

-- Screen #47 --

Q200

**To finish the study, please complete the follow-up survey questions on the next three pages.**

**Please answer these questions carefully and honestly.**

→

-- Screen #48 --

QID145

**Follow-up survey (Page 1 out of 3)**

Q1

Please rate your agreement with the following statements about your local community pharmacist:

| | Strongly Disagree | Disagree | Neutral | Agree | Strongly Agree | No basis for observation |
|---|---|---|---|---|---|---|
| The pharmacist provides kindness/positive intention. | ○ | ○ | ○ | ○ | ○ | ○ |
| The pharmacist is competent. | ○ | ○ | ○ | ○ | ○ | ○ |
| The pharmacist works with integrity. | ○ | ○ | ○ | ○ | ○ | ○ |
| The pharmacist makes me feel at ease. | ○ | ○ | ○ | ○ | ○ | ○ |
| The pharmacist is fair. | ○ | ○ | ○ | ○ | ○ | ○ |
| The pharmacist communicates well | ○ | ○ | ○ | ○ | ○ | ○ |
| The pharmacist is dependable. | ○ | ○ | ○ | ○ | ○ | ○ |
| The pharmacist is confidential. | ○ | ○ | ○ | ○ | ○ | ○ |
| I trust the pharmacist | ○ | ○ | ○ | ○ | ○ | ○ |

▸ *Response required.*

▸ *Hidden page timer (Q164) records time spent on this page.*

→

-- Screen #49 --

Q165

**Follow-up survey (Page 2 out of 3)**

38.

What is the approximate distance from your home to the nearest pharmacy? (miles)

- Less than 1
- 2 - 5
- 6 - 10
- Greater than 10

*Response required.*

39.

When was your last visit to a pharmacy?

- Within 3 months
- 3 - 6 months
- Greater than 6 months

*Response required.*

40.

Do you have a diagnosed chronic medical condition (defined here as a condition requiring medication to be routinely refilled)?

- Yes
- No

*Response required.*

*Hidden page timer (Q166) records time spent on this page.*

→

-- Screen #50 --

Q201

**Follow-up survey (Page 3 out of 3)**

41.

Please indicate how much you agree with the following statements.

| | Strongly Disagree | Disagree | Neutral | Agree | Strongly Agree |
|---|---|---|---|---|---|
| I made each decision in this study carefully. | ○ | ○ | ○ | ○ | ○ |
| I made decisions in this study randomly. | ○ | ○ | ○ | ○ | ○ |
| I understood what my decisions meant for how much a patient may pay. | ○ | ○ | ○ | ○ | ○ |
| Select the button that is furthest to the left. | ○ | ○ | ○ | ○ | ○ |
| Select the button that is furthest to the right. | ○ | ○ | ○ | ○ | ○ |

‣ *Response required.*

gender

What is your stated gender?

- ○ Male
- ○ Female
- ○ Non-binary

‣ *Response required.*

race

What is your stated race? (Select all that apply.)

- ☐ Caucasian or white
- ☐ Hispanic
- ☐ African American or Black
- ☐ Asian or Pacific Islander
- ☐ Native American
- ☐ Other

‣ *Response required.*

option_text

(Optional): If you would like to, please describe how you made decisions in this study.

*Hidden page timer (Q205) records time spent on this page.*

→

End of survey: participants are redirected to https://ucsd.co1.qualtrics.com/jfe/form/SV_6zXKN99kyr14PEa.

**Supplement Table 2. Comparison of survey to U.S. Census Bureau data for annual household income**[41]

| Annual household income | Survey | U.S. households, 2024 ACS 5-year | Difference |
|---|---|---|---|
| Below $30,000 | 18.8% | 17.9% | +0.9 pp |
| $30,000–$74,999 | 36.8% | 29.0% | +7.8 pp |
| $75,000–$149,999 | 32.3% | 30.2% | +2.1 pp |
| $150,000 or more | 12.2% | 23.0% | −10.8 pp |

National percentages calculated by combining the income brackets in ACS Table B19001. Differences use unrounded percentages; pp = percentage points.

**Supplement Table 3. Comparison of geographic distribution between survey and U.S. Census Bureau data**[42,43]

| U.S. region | Survey | U.S. adult population, 2025 | Difference |
|---|---|---|---|
| Northeast | 18.2% | 17.5% | +0.7 |
| Midwest | 19.4% | 21.3% | −1.9 |
| South | 41.6% | 39.2% | +2.4 |
| West | 20.8% | 22.0% | −1.2 |

Survey percentages recalculated from respondent counts; national percentages calculated from 2025 Census regional population estimates after excluding residents younger than 18.

**Supplement Table 4. Demographic comparison of survey to U.S. Census Bureau data[43]**

| Characteristic | Survey | U.S. adults 18+ | Assessment |
|---|---|---|---|
| Male | 48% | ~49% | Closely aligned |
| Female | 50% | ~51% | Closely aligned |
| Nonbinary | 2% | Not directly comparable | Different measurement definitions |
| Ages 18–26 | 15% | ~16% | Approximately aligned |
| Ages 27–34 | 14% | ~14% | Approximately aligned |
| Ages 35–49 | 25% | ~25% | Approximately aligned |
| Ages 50–67 | 36% | ~29% | Overrepresented |
| Over 67 | 10% | ~16% | Underrepresented |
| Hispanic | 8% | ~17% | Substantially underrepresented |
| Black/African American | 12% | ~12–13% | Approximately aligned |
| Asian/Pacific Islander | 6% | ~6–7% | Approximately aligned |
| Multiple races | 8% | Definition-dependent | Cannot establish equivalence |

Age estimates are approximations based on Census age distributions, including the 2024 ACS and 2025 population estimates.
